# A Metabolic-Inflammatory Phenotype of Pelvic Floor Dysfunction: A Machine Learning-Based Discovery in a Nationally Representative U.S. Cohort

**DOI:** 10.1101/2025.10.29.25339110

**Authors:** Jingming Yang, Duo Zhao, Lan Wang

**Affiliations:** Department of Urology, Huai’an Traditional Chinese Medicine Hospital, Huai’an, Jiangsu, China; Department of Orthopedics, The First People’s Hospital of Guannan: Lianyungang, Lianyungang, Jiangsu, China; Department of Obstetrics and Gynecology, Huai’an 82 hospital, China RongTong Medical Healthcare Group Co. Ltd, Huai’an, Jiangsu, China

**Keywords:** Pelvic Floor Dysfunction, Machine Learning, Clinical Subtypes / Phenotyping, Metabolic Syndrome, NHANES, Urogynecology

## Abstract

**Background:** Pelvic floor dysfunction (PFD) is a highly prevalent and heterogeneous condition among women. The traditional view of PFD as a single clinical entity limits the understanding of its complex pathophysiology and hinders the development of personalized therapies.

**Objective:** We aimed to deconstruct the heterogeneity of PFD by using unsupervised machine learning to identify distinct clinical subtypes in a nationally representative sample of U.S. women.

**Methods:** This cross-sectional study included 7,291 female participants aged ≥20 from the National Health and Nutrition Examination Survey (NHANES) 2005-2012. We employed a two-stage analytical approach. First, K-means clustering was performed on PFD-positive women using 12 physiological features to identify subtypes. Second, we developed and validated seven multiclass machine learning models to predict subtype membership (Healthy Control, Phenotype 1, Phenotype 2). The best model was interpreted using SHAP (SHapley Additive exPlanations).

**Results:** Two distinct clinical subtypes were identified among PFD-positive women. Phenotype 1 (Metabolic-Inflammatory) was characterized by severe metabolic disturbances, including central obesity (mean BMI 36.1 kg/m²), and high prevalence of diabetes (21%) and hypertension (50%). In contrast, Phenotype 2 (Metabolically-Healthy) exhibited PFD symptoms but had a metabolic profile nearly identical to healthy controls (mean BMI 25.4 kg/m²; diabetes prevalence 3.7%). Our prediction models accurately distinguished the three groups, with a Neural Network achieving the highest macro-AUC of 0.848. SHAP analysis revealed that waist circumference was overwhelmingly the most important predictor for differentiating the subtypes.

**Conclusions:** PFD is not a monolithic disorder but comprises at least two distinct clinical subtypes. One subtype is intrinsically linked to systemic metabolic and inflammatory disease, while the other appears to be driven by more traditional risk factors. This novel, data-driven classification provides a new framework for understanding PFD and may pave the way for precision diagnostics and targeted therapeutic strategies.

## 1. Introduction

Pelvic floor dysfunction (PFD) encompasses a group of highly prevalent yet under-recognized clinical syndromes affecting hundreds of millions of women worldwide, primarily manifesting as stress urinary incontinence (SUI), urgency urinary incontinence (UUI), pelvic organ prolapse (POP), and fecal incontinence (FI) (1–3).The impact of PFD on women’s quality of life is profound; it not only causes significant physical distress but also imposes substantial psychological and economic burdens (4–6).Furthermore, among middle-aged and older women, symptoms such as urgency urinary incontinence significantly elevate the risk of falls, posing a serious health threat (7).As the global population ages, the prevalence of PFD is projected to rise, establishing it as an escalating public health challenge.

While conventional epidemiological research has identified several key risk factors for PFD—with advancing age, parity (particularly vaginal delivery), menopause, and obesity being the most recognized drivers (6,8–10)—a persistent challenge for both clinicians and researchers is the condition’s marked clinical heterogeneity (1,11).Individuals with similar risk exposures often exhibit wide variations in symptom presentation, disease progression, and therapeutic response (11).Current clinical practice and research frequently treat PFD as a single, homogenous entity. This monolithic view critically limits our understanding of its complex pathophysiology and impedes the development of personalized prevention and treatment strategies (12–14).

In recent years, machine learning, particularly unsupervised clustering, has emerged as a powerful data-driven tool for deconstructing complex, heterogeneous diseases, successfully identifying clinically meaningful subtypes in fields such as oncology, diabetes, and cardiovascular disease (15–19).This approach, which is not reliant on a priori assumptions, can objectively uncover latent patient clusters from high-dimensional data, thereby challenging traditional medical paradigms and advancing the stratification required for precision medicine (19,20). We hypothesized that PFD is not a single pathological entity but rather a collection of distinct subtypes driven by different pathophysiological pathways. Specifically, we posited that in addition to the classic subtype—primarily driven by mechanical injury and tissue aging—another subtype exists that is intrinsically linked to systemic metabolic dysregulation and chronic low-grade inflammation, which has not yet been clearly defined.

Accordingly, this study aimed to systematically deconstruct the heterogeneity of female PFD by applying unsupervised machine learning for the first time to a large, nationally representative dataset from the U.S. National Health and Nutrition Examination Survey (NHANES). Our specific objectives were to: 1) objectively identify data-driven clinical subtypes within the PFD-positive population; 2) comprehensively characterize and compare these subtypes across multiple domains, including demographics, clinical comorbidities, and metabolic, inflammatory, and functional status; and 3) develop and validate a machine learning model to accurately predict subtype membership and identify the most critical clinical predictors. Through this research, we expect to provide new insights into the etiology of PFD and to lay the groundwork for future precision diagnostics and subtype-targeted therapeutic strategies.

## 2. Methods

### 2.1 Study Design and Population

The authors accessed the U.S. National Health and Nutrition Examination Survey (NHANES) 2005-2012 data for research purposes on September 20, 2025.The authors did not have access to information that could identify individual participants during or after data collection, as the dataset is fully de-identified. This study was a secondary analysis of data from the NHANES. The NHANES protocol was approved by the National Center for Health Statistics (NCHS) Research Ethics Review Board, and all participants provided written informed consent. As this research utilized publicly available, de-identified data, it was determined to be exempt from review by our institutional review board. The participant selection process is detailed in Figure 1. We began with 40,790 non-pregnant individuals from the NHANES 2005–2012 cycles, from which we identified 20,533 female participants. We then restricted the cohort to adults aged 20 years and older, resulting in 11,547 eligible individuals. To focus on PFD driven by non-iatrogenic factors, we subsequently excluded participants with missing PFD outcome data (N=1,777), a history of major pelvic surgery (N=2,121), specific neurological diseases (including stroke, multiple sclerosis, spinal cord injury, and Parkinson’s disease; N=193), and cancers with a direct pelvic impact (including gynecological, colorectal, or urological cancers; N=165). Following this rigorous screening process, a final analytical cohort of 7,291 participants was established.To further validate the biological characteristics of the identified subtypes, we also conducted sensitivity analyses in available data subsets using an objective physiological measure (grip strength) and a marker of systemic inflammation (periodontitis) (see Supplementary Material 1).

**Figure 1.**
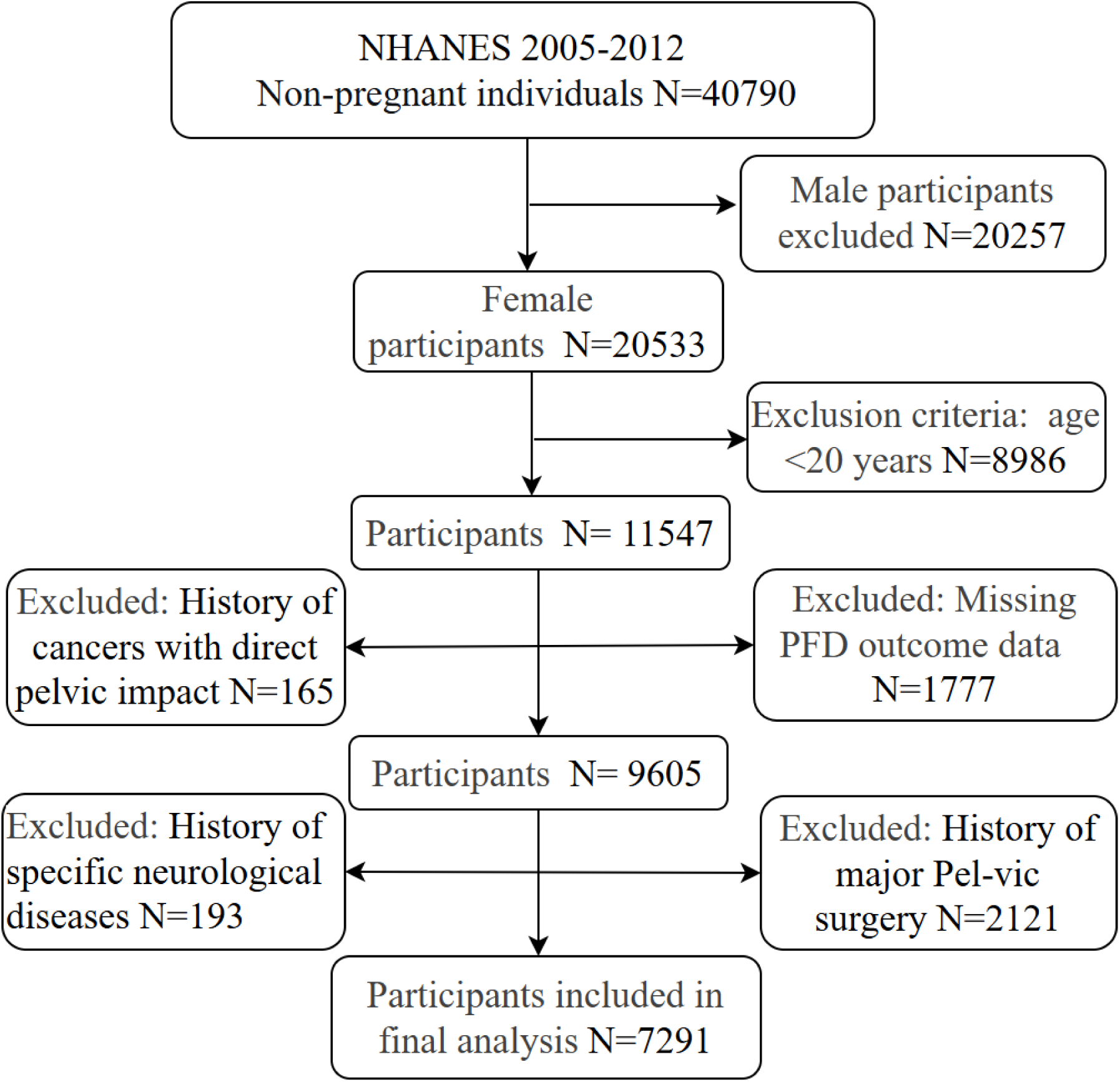
Flowchart of participants selection.

### 2.2 Variable Definitions

Pelvic floor dysfunction (PFD) was defined by the presence of one or more of the following symptoms: stress urinary incontinence (SUI), urgency urinary incontinence (UUI), fecal incontinence (FI), or pelvic organ prolapse (POP). Symptom presence was determined using standardized questions from the NHANES questionnaires. Detailed definitions for all outcome variables and covariates are provided in Supplementary Material 2.

### 2.3 Statistical Analysis

All data cleaning, management, and analyses were performed using R software (version 4.4.2), incorporating the survey package to account for the complex, multistage sampling design of NHANES (weighting, strata, and clusters). All statistical tests were two-sided, and a *P*-value < 0.05 was considered statistically significant. Regarding the handling of missing data, we first assessed the proportion of missingness for all covariates. Any covariate with a missing rate exceeding 20% was considered unreliable and was, in principle, excluded from the final analytical models, rather than excluding the participants. Missing data in covariates were addressed using multiple imputation by chained equations (MICE), a highly recommended method in current literature (R package mice, m=20 imputations) (21,22). Our analytical workflow consisted of two main stages: Stage 1: Unsupervised Discovery of PFD Subtypes. First, we applied the K-means clustering algorithm to the PFD-positive cohort to objectively identify latent clinical subtypes. The clustering was based on 12 carefully selected core physiological features spanning the domains of obesity, metabolism, nutrition, physical function, and mental health. The optimal number of clusters (K) was determined by the integrated use of the elbow and silhouette methods. Stage 2: Supervised Prediction and Interpretation of Subtypes. After identifying two clinical subtypes, we categorized the study population into three classes: Healthy Controls, Phenotype 1, and Phenotype 2. The dataset was then randomly partitioned into a training set (80%) and an independent test set (20%). On the training set, we developed and compared seven distinct machine learning algorithms—including neural networks, XGBoost, logistic regression, random forest, support vector machines, naive Bayes, and K-nearest neighbors—to build multiclass models capable of accurately predicting an individual’s class membership. Model performance was evaluated on the test set using the macro-average area under the receiver operating characteristic curve (AUC) as the primary metric. To interpret the best-performing model and identify the key drivers of subtype differentiation, we employed SHAP (SHapley Additive exPlanations) analysis to quantify the contribution of each predictor.Baseline characteristics were described using weighted means (standard deviations) or medians (interquartile ranges) for continuous variables and weighted percentages for categorical variables. Group comparisons were conducted using design-based t-tests (or Kruskal-Wallis tests) and chi-square tests with the Rao & Scott adjustment, as appropriate. We also performed a sensitivity analysis by constructing three nested predictive models to assess the change in the relative importance of parity across varying levels of model complexity.

## 3. Results

### 3.1 Participant Baseline Characteristics

The final analytical cohort included 7,291 participants, representing 72,325,873 U.S. adult women. Of these, 36,483,067 (a weighted 50.5%) were identified as having at least one type of pelvic floor dysfunction (the PFD-positive group), while 35,842,806 (a weighted 49.5%) had no PFD symptoms (the PFD-negative group). The baseline characteristics of these two groups are detailed in Table 1.

**Table 1.**
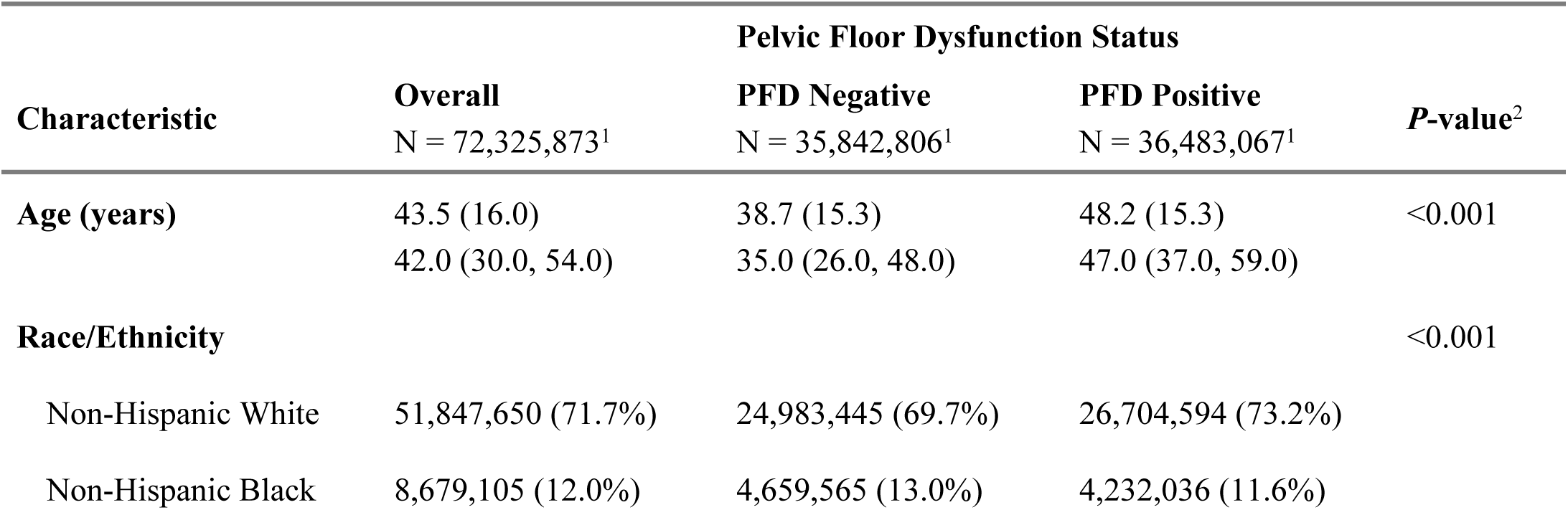
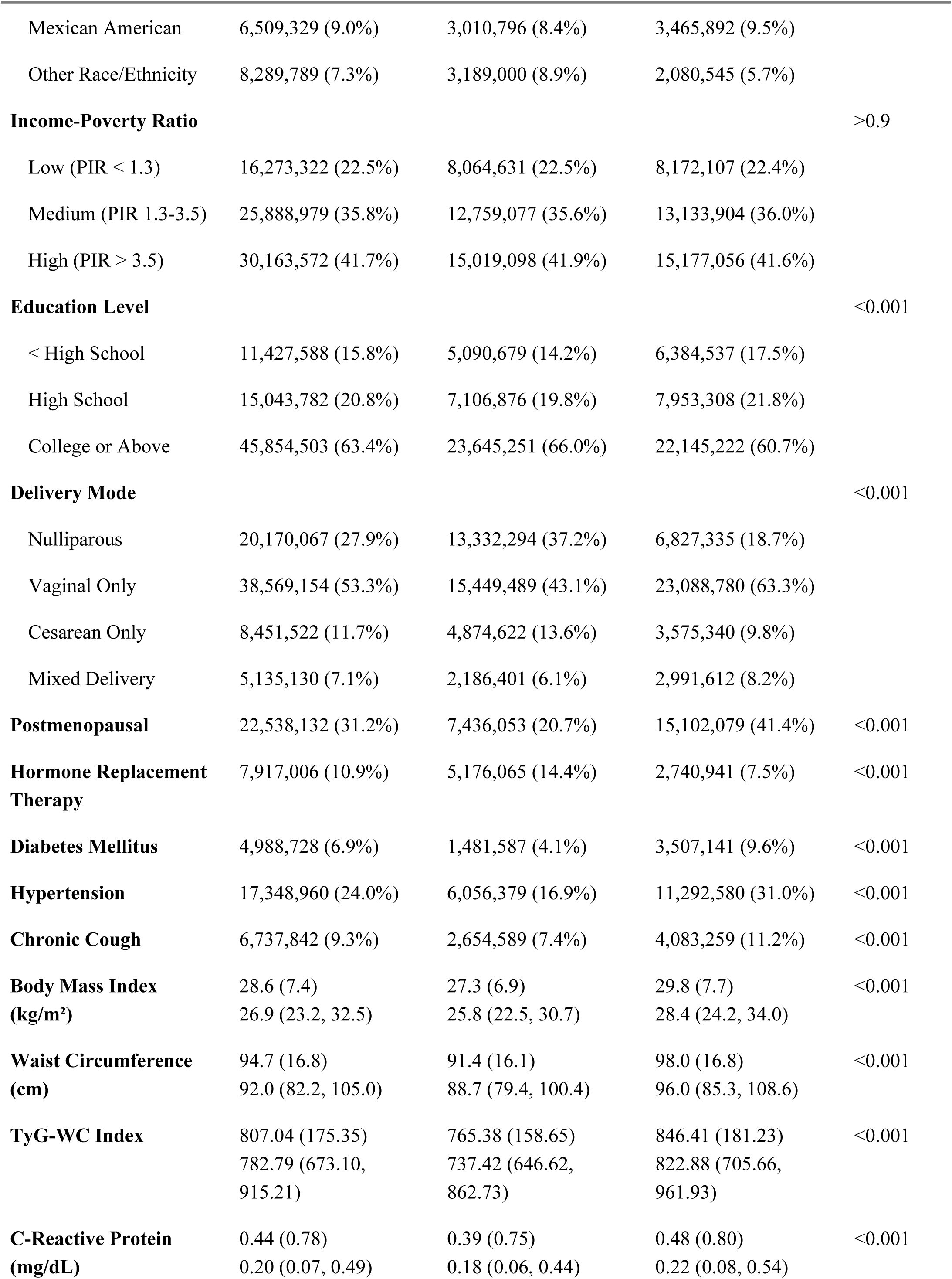
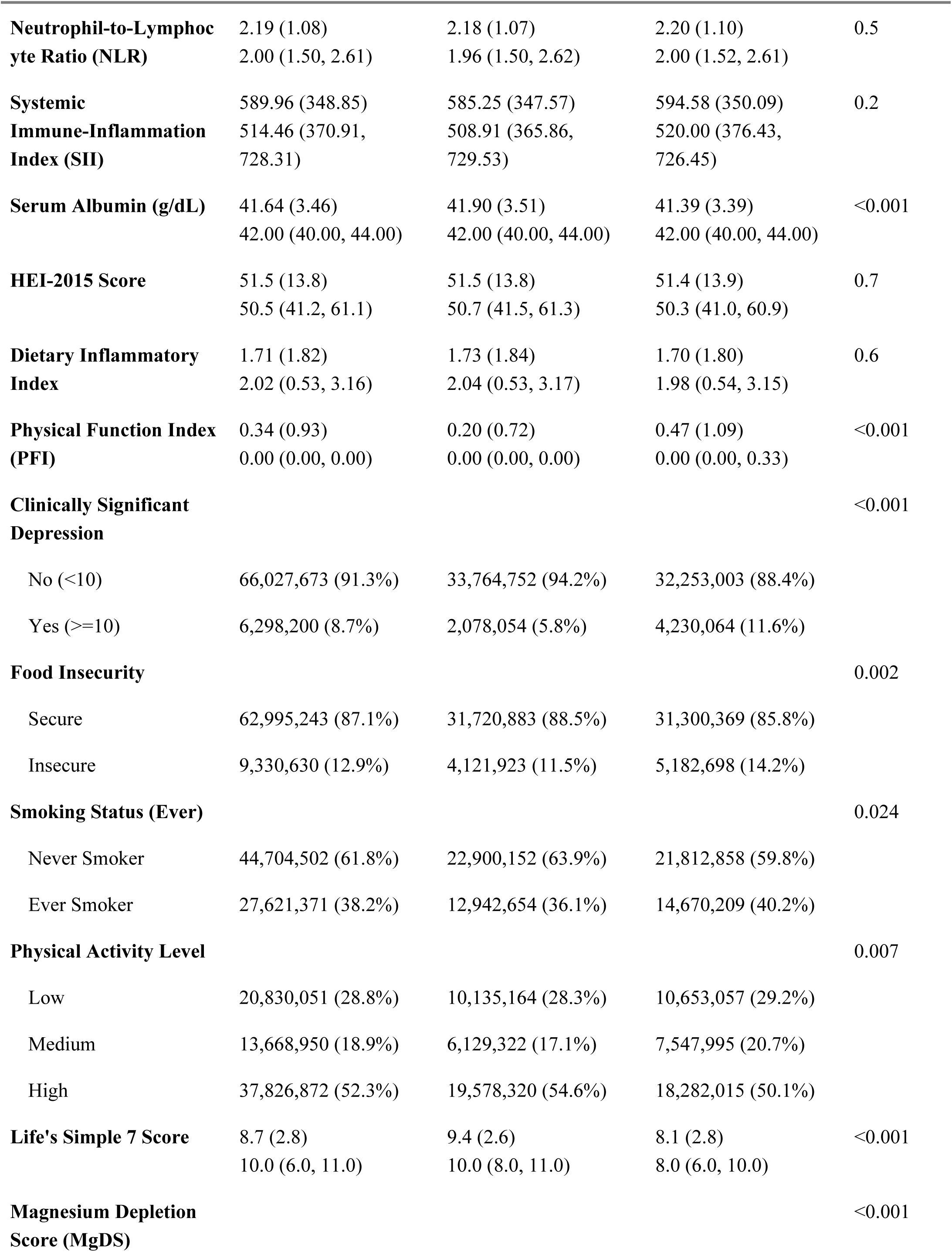
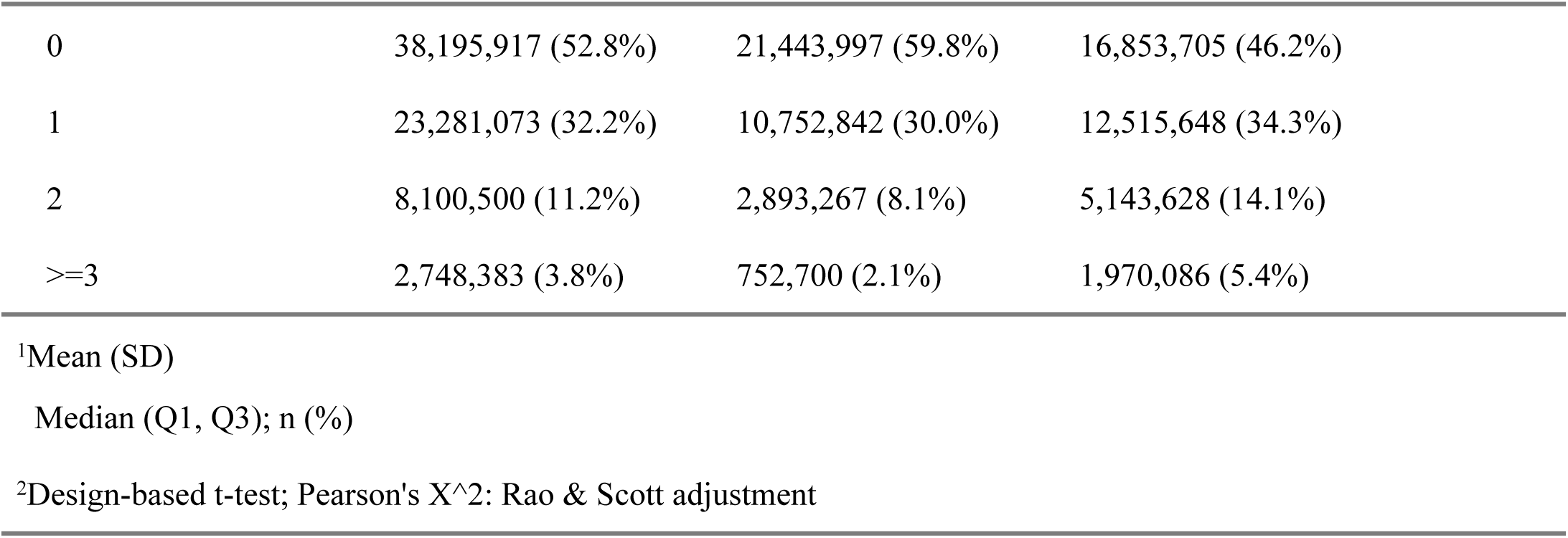
Baseline Characteristics of the Study Population, Stratified by Pelvic Floor Dysfunction Status.

Compared to their PFD-negative counterparts, women in the PFD-positive group were significantly older (mean age: 48.2 vs. 38.7 years, *P* < 0.001). Regarding reproductive history, the PFD-positive group had a significantly lower proportion of nulliparous women (18.7% vs. 37.2%) and a significantly higher proportion who had experienced only vaginal deliveries (63.3% vs. 43.1%; *P* < 0.001). Furthermore, the prevalence of postmenopausal status was nearly double in the PFD-positive group (41.4% vs. 20.7%, *P* < 0.001).

In terms of comorbidities, the prevalence of several chronic conditions was significantly higher in the PFD-positive group, including diabetes (9.6% vs. 4.1%), hypertension (31.0% vs. 16.9%), and chronic cough (11.2% vs. 7.4%) (all *P* < 0.001).

Markers of central obesity and metabolic dysregulation were also more pronounced in the PFD-positive group. Their mean body mass index (BMI) (29.8 vs. 27.3 kg/m²), waist circumference (98.0 vs. 91.4 cm), and TyG-WC index—an indicator of insulin resistance and central obesity—were all significantly higher than in the PFD-negative group (all *P* < 0.001). Regarding inflammatory markers, C-reactive protein (CRP) and serum albumin levels showed statistically significant differences (both *P* < 0.001), whereas the neutrophil-to-lymphocyte ratio (NLR) and systemic immune-inflammation index (SII) did not differ significantly between the groups.

In the domains of functional, lifestyle, and psychosocial factors, the PFD-positive group reported poorer physical function (higher PFI scores), a higher prevalence of clinical depressive symptoms (12% vs. 5.8%), a greater proportion experiencing food insecurity (14.2% vs. 11.5%), and a higher rate of ever-smokers (40.2% vs. 36.1%) (all *P* < 0.05). The Life’s Simple 7 (LS7) score, representing overall cardiovascular health, was significantly lower in the PFD-positive group (mean: 8.1 vs. 9.4, *P* < 0.001), while the Magnesium Depletion Score (MgDS) indicated a more severe risk of magnesium deficiency in this group (*P* < 0.001).

Notably, no statistically significant differences were observed between the two groups in terms of the poverty-income ratio (PIR), the Healthy Eating Index (HEI-2015), or the Dietary Inflammatory Index (DII).

### 3.2 Identification and Definition of PFD Clinical Subtypes

To investigate the heterogeneity within the PFD-positive population, we applied K-means clustering, an unsupervised learning algorithm, to 12 core physiological features. The optimal number of clusters (K) was determined using both the elbow method and the silhouette method. The elbow method revealed a sharp decrease in the within-cluster sum of squares (WSS) as K increased from 1 to 2, indicating a potential inflection point at K=2 (Supplementary Material 3, Figure S1). More definitively, the silhouette method showed that the average silhouette width peaked at K=2 (approx. 0.24) before declining with additional clusters, providing strong evidence for selecting two clusters (Supplementary Material 3, Figure S2). Consequently, we classified the PFD-positive population into two distinct clinical subtypes, designated as Phenotype 1 and Phenotype 2.

Table 2 provides a detailed comparison of baseline characteristics among the Healthy Control group, Phenotype 1, and Phenotype 2. The results demonstrate that these two subtypes possess strikingly different clinical and biological profiles.

**Table 2.** Baseline Characteristics of Healthy Controls and the Two Identified PFD Phenotypes.

| Characteristic | Participant Group |  |  | P-value<sup>2</sup> |
| --- | --- | --- | --- | --- |
|  | Healthy Control<br>N = 35,842,806<sup>1</sup> | Phenotype 1<br>N = 6,386,347<sup>1</sup> | Phenotype 2<br>N = 10,111,038<sup>1</sup> |  |
| Age (years) | 38.7 (15.3) | 50.1 (15.0) | 46.9 (15.0) | <0.001 |
| Race/Ethnicity |  |  |  | <0.001 |
| Non-Hispanic White | 24,983,445 (69.7%) | 4,495,988 (70.4%) | 7,583,279 (75.0%) |  |
| Non-Hispanic Black | 4,659,565 (13.0%) | 894,089 (14.0%) | 909,993 (9.0%) |  |
| Mexican American | 3,010,796 (8.4%) | 702,498 (11.0%) | 889,771 (8.8%) |  |
| Other Race/Ethnicity | 3,189,000 (8.9%) | 293,772 (4.6%) | 727,995 (7.2%) |  |

| Characteristic | Participant Group |  |  | *P*-value<sup>2</sup> |
| --- | --- | --- | --- | --- |
|  | Healthy Control<br>N = 35,842,806<sup>1</sup> | Phenotype 1<br>N = 6,386,347<sup>1</sup> | Phenotype 2<br>N = 10,111,038<sup>1</sup> |  |
| **Income-Poverty Ratio** |  |  |  | <0.001 |
| Low (PIR < 1.3) | 8,064,631 (22.5%) | 1,877,690 (29.4%) | 1,739,100 (17.2%) |  |
| Medium (PIR 1.3-3.5) | 12,759,077 (35.6%) | 2,624,688 (41.1%) | 3,639,974 (36.0%) |  |
| High (PIR > 3.5) | 15,019,098 (41.9%) | 1,883,347 (29.5%) | 4,731,964 (46.8%) |  |
| **Education Level** |  |  |  | <0.001 |
| < High School | 5,090,679 (14.2%) | 1,390,233 (21.8%) | 1,445,878 (14.3%) |  |
| High School | 7,106,876 (19.8%) | 1,532,723 (24.0%) | 1,981,763 (19.6%) |  |
| College or Above | 23,645,251 (66.0%) | 3,463,391 (54.2%) | 6,683,397 (66.1%) |  |
| **Delivery Mode** |  |  |  | <0.001 |
| Nulliparous | 13,332,294 (37.2%) | 951,757 (14.9%) | 1,910,985 (18.9%) |  |
| Vaginal Only | 15,449,489 (43.1%) | 4,158,281 (65.1%) | 6,582,287 (64.1%) |  |
| Cesarean Only | 4,874,622 (13.6%) | 695,112 (10.9%) | 909,993 (9.0%) |  |
| Mixed Delivery | 2,186,401 (6.1%) | 581,197 (9.1%) | 707,773 (7.0%) |  |
| **Postmenopausal** | 7,436,053 (20.7%) | 3,080,315 (48.2%) | 3,678,578 (36.4%) | <0.001 |
| **Hormone Replacement Therapy** | 5,176,065 (14.4%) | 341,674 (5.4%) | 927,332 (9.2%) | <0.001 |
| **Diabetes Mellitus** | 1,481,587 (4.1%) | 1,336,263 (20.9%) | 371,843 (3.7%) | <0.001 |
| **Hypertension** | 6,056,379 (16.9%) | 3,189,821 (49.9%) | 2,055,916 (20.3%) | <0.001 |
| **Chronic Cough** | 2,654,589 (7.4%) | 956,415 (15.0%) | 774,158 (7.7%) | <0.001 |
| **Body Mass Index (kg/m<sup>2</sup>)** | 27.3 (6.9) | 36.1 (6.5) | 25.4 (4.1) | <0.001 |
| **Waist Circumference (cm)** | 91.4 (16.1) | 113.5 (12.7) | 88.0 (9.6) | <0.001 |
| **TyG-WC Index** | 765.38 (158.65) | 1,025.40 (132.81) | 733.36 (97.08) | <0.001 |
| **C-Reactive Protein (mg/dL)** | 0.39 (0.75) | 0.73 (0.92) | 0.30 (0.63) | <0.001 |
| **Neutrophil-to-Lymphocyte Ratio (NLR)** | 2.18 (1.07) | 2.25 (1.04) | 2.16 (1.07) | 0.346 |
| **Systemic Immune-Inflammation Index (SII)** | 585.25 (347.57) | 613.89 (312.28) | 558.56 (301.32) | 0.026 |
|  | Healthy Control<br>N = 35,842,806<sup>1</sup> | Phenotype 1<br>N = 6,386,347<sup>1</sup> | Phenotype 2<br>N = 10,111,038<sup>1</sup> |  |
| **Serum Albumin (g/dL)** | 41.90 (3.51) | 39.86 (3.49) | 42.03 (2.97) | <0.001 |
| **HEI-2015 Score** | 51.5 (13.8) | 49.4 (12.9) | 52.8 (14.5) | 0.008 |
| **Dietary Inflammatory Index** | 1.73 (1.84) | 1.85 (1.76) | 1.55 (1.91) | 0.059 |
| **Physical Function Index (PFI)** | 0.20 (0.72) | 0.78 (1.35) | 0.20 (0.60) | <0.001 |
| **Clinically Significant Depression** |  |  |  | <0.001 |
| No (<10) | 33,764,752 (94.2%) | 5,287,495 (82.8%) | 9,504,376 (94.0%) |  |
| Yes (≥10) | 2,078,054 (5.8%) | 1,098,852 (17.2%) | 606,662 (6.0%) |  |
| **Food Insecurity** |  |  |  | <0.001 |
| Secure | 31,720,883 (88.5%) | 5,097,305 (79.8%) | 9,059,490 (89.6%) |  |
| Insecure | 4,121,923 (11.5%) | 1,289,042 (20.2%) | 1,051,548 (10.4%) |  |
| **Smoking Status (Ever)** |  |  |  | 0.071 |
| Never Smoker | 22,900,152 (63.9%) | 3,653,034 (57.2%) | 6,309,153 (62.4%) |  |
| Ever Smoker | 12,942,654 (36.1%) | 2,733,313 (42.8%) | 3,801,885 (37.6%) |  |
| **Physical Activity Level** |  |  |  | 0.386 |
| Low | 10,135,164 (28.3%) | 1,883,969 (29.5%) | 2,649,092 (26.2%) |  |
| Medium | 6,129,322 (17.1%) | 1,264,497 (19.8%) | 2,022,208 (20.0%) |  |
| High | 19,578,320 (54.6%) | 3,237,881 (50.7%) | 5,439,738 (53.8%) |  |
| **Life's Simple 7 Score** | 9.4 (2.6) | 6.3 (2.1) | 9.7 (2.2) | <0.001 |
| **Magnesium Depletion Score (MgDS)** |  |  |  | <0.001 |
| 0 | 21,443,997 (59.8%) | 2,816,648 (44.1%) | 4,984,804 (49.3%) |  |
| 1 | 10,752,842 (30.0%) | 2,183,901 (34.2%) | 3,508,468 (34.7%) |  |
| 2 | 2,893,267 (8.1%) | 932,407 (14.6%) | 1,263,880 (12.5%) |  |
| ≥3 | 752,700 (2.1%) | 453,391 (7.1%) | 353,886 (3.5%) |  |

<sup>1</sup>Mean (SD); n (%)
<sup>2</sup>Design-based KruskalWallis test; Pearson's X<sup>2</sup>: Rao & Scott adjustment

Phenotype 1 was defined as the “Metabolic-Inflammatory Phenotype,” representing 6,386,347 women. It was distinguished by severe metabolic dysregulation and systemic inflammation. Compared to healthy controls, Phenotype 1 exhibited a dramatically higher prevalence of diabetes (20.9% vs. 4.1%) and hypertension (49.9% vs. 16.9%). These individuals presented with severe obesity, having a mean BMI of 36.1 kg/m² and a mean waist circumference of 113.5 cm, and their TyG-WC index, an indicator of insulin resistance, was extremely high (mean 1025.40). Regarding inflammatory markers, this subtype showed significantly elevated C-reactive protein (CRP) levels (mean 0.73 mg/dL) and reduced serum albumin (mean 39.86 g/dL). On the functional and psychosocial level, Phenotype 1 reported poorer physical function (PFI score 0.78), a much higher prevalence of clinical depressive symptoms (17.2% vs. 5.8%), greater food insecurity (20.2% vs. 11.5%), and exceptionally poor cardiovascular health (mean LS7 score of only 6.3). Phenotype 2, in contrast, was defined as the “Metabolically-Healthy Phenotype,” representing 10,111,038 women. This subtype’s profile was in stark contrast to that of Phenotype 1. Despite also having PFD, their metabolic parameters were nearly indistinguishable from the healthy control group, with a mean BMI of 25.4 kg/m², a mean waist circumference of 88.0 cm, a diabetes prevalence of only 3.7%, and a hypertension prevalence of 20%. Their CRP level (0.30 mg/dL) was also within the normal range. Notably, this subtype’s cardiovascular health score was even slightly higher than that of the healthy controls (mean LS7 score: 9.7 vs. 9.4). However, compared to the healthy control group, these women were older (46.9 vs. 38.7 years) and had a similarly high proportion of vaginal deliveries (64.1% vs. 43.1%).

Table 2 represents a subset of the participants from Table 1; see Supplementary Material 2 for a detailed explanation.

### 3.3 Performance of Multiclass Predictive Models

To develop a model capable of accurately distinguishing among the three clinical states (Healthy Control, Phenotype 1, Phenotype 2), we trained and evaluated seven different machine learning algorithms. Model performance was assessed on an independent test set and quantified using the macro-average area under the receiver operating characteristic (ROC) curve (AUC).Figure 2 illustrates the ROC curves for all models on the test set. All algorithms demonstrated strong predictive capabilities, with AUC values substantially exceeding the 0.5 threshold of random chance. The neural network model performed best, achieving the highest macro-average AUC of 0.848. The performance of several other models was highly competitive and nearly equivalent. Specifically, XGBoost (AUC = 0.837), logistic regression (AUC = 0.834), and random forest (AUC = 0.831) all showed robust discriminatory power. Support vector machine (AUC = 0.825) and naive Bayes (AUC = 0.792) followed, while the K-nearest neighbors algorithm was the weakest performer for this task (AUC = 0.768). Overall, multiple models successfully achieved effective prediction of the clinical subtypes, with AUCs generally above 0.83, indicating that our identified PFD subtypes are highly distinguishable.

**Figure 2.**
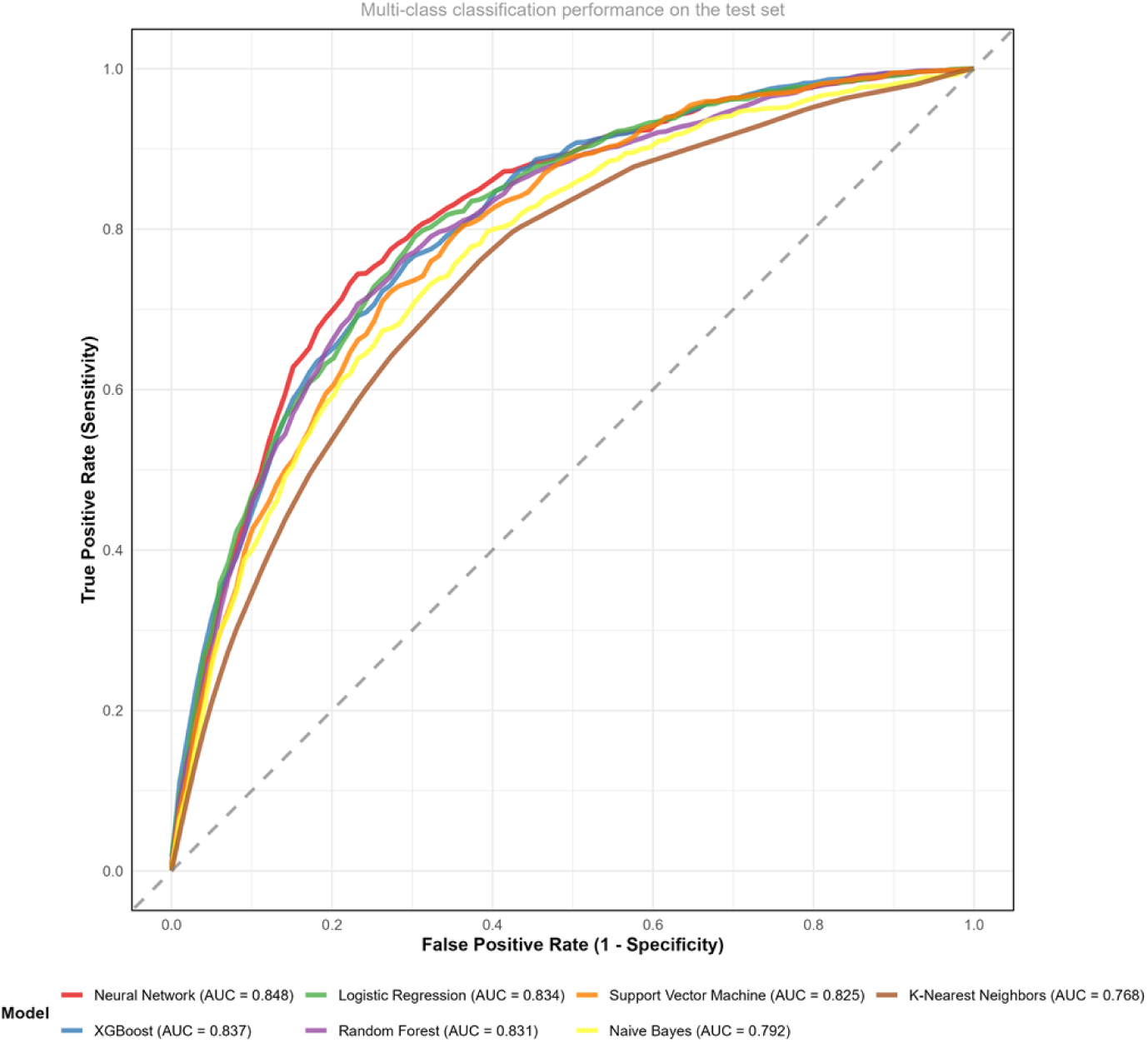
ROC curves comparison of machine learning models.

### 3.4 Key Predictors for Differentiating PFD Subtypes

To identify the most critical factors for distinguishing between the two PFD clinical subtypes, we analyzed the best-performing and interpretable XGBoost model. We employed SHAP (SHapley Additive exPlanations) values to quantify the contribution of each feature. Figure 3 displays the top ten predictors ranked by their feature importance.

**Figure 3.**
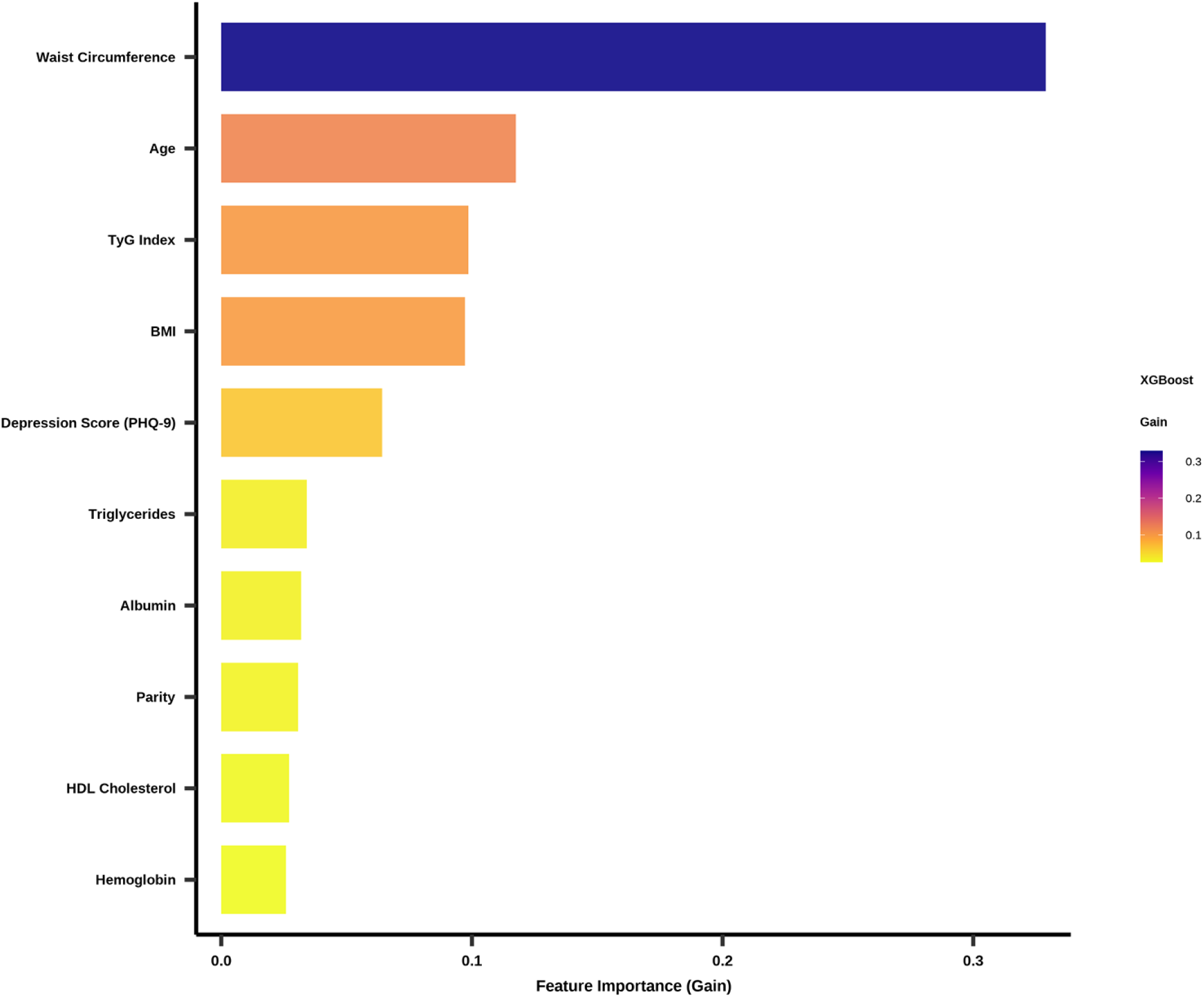
Top 10 predictive features (XGBoost).

The analysis revealed that waist circumference emerged as the single most dominant predictor, with an importance score exceeding 0.3—nearly three times that of the second-ranked feature, age. This factor played a decisive role in subtype differentiation.

Beyond waist circumference, other key predictors clustered into three primary domains: 1) Demographic and Core Metabolic Metrics: Age (importance score: 0.117) was the second most important factor, closely followed by the TyG index (0.099), an indicator of insulin resistance, and Body Mass Index (BMI, 0.097). 2)Systemic Health and Psychological Status: The depression score (PHQ-9) also demonstrated a significant contribution.3)Biological Markers and Reproductive History: The remaining top-ten factors included lipid profile markers (triglycerides, HDL cholesterol), serum albumin (reflecting nutritional and inflammatory status), parity (a classic obstetric risk factor), and hemoglobin level (an indicator of overall health).

In summary, the feature importance analysis demonstrates that metrics of metabolic dysregulation centered on central obesity are the primary drivers for differentiating between the two PFD subtypes. These are followed by age and composite indicators reflecting systemic health and psychological status.

### 3.5 Sensitivity Analysis: Predictive Importance of Parity in Nested Models

To investigate the role of parity, a traditional risk factor, across predictive models of varying complexity, we conducted a sensitivity analysis. We constructed three nested models: 1) a “basic clinical model” with 5 core variables; 2) a “metabolic model” incorporating 10 variables; and 3) a “systemic health model” with 13 variables. Detailed results are available in Supplementary Material 4.

As shown in Figure 4, the relative importance of parity systematically decreased as model complexity increased. In the basic clinical model, which included only BMI, age, parity, race/ethnicity, and education, parity ranked as the third most important feature, following BMI and age, with an importance score of 8.81 (compared to 100 for BMI and 47.24 for age). When metabolic indicators were added (10 features in total), the rank of parity dropped to 5th. In the final, most comprehensive model that included systemic health indicators (13 features in total), its rank further declined to 6th.This finding indicates that while parity serves as a significant independent predictor in a simple clinical assessment, a substantial portion of its predictive value is explained or absorbed by more comprehensive metabolic and systemic health metrics. This strongly suggests that the long-term association of parity with PFD may be primarily mediated through its complex interplay with a woman’s current metabolic and systemic health status.

**Figure 4.**
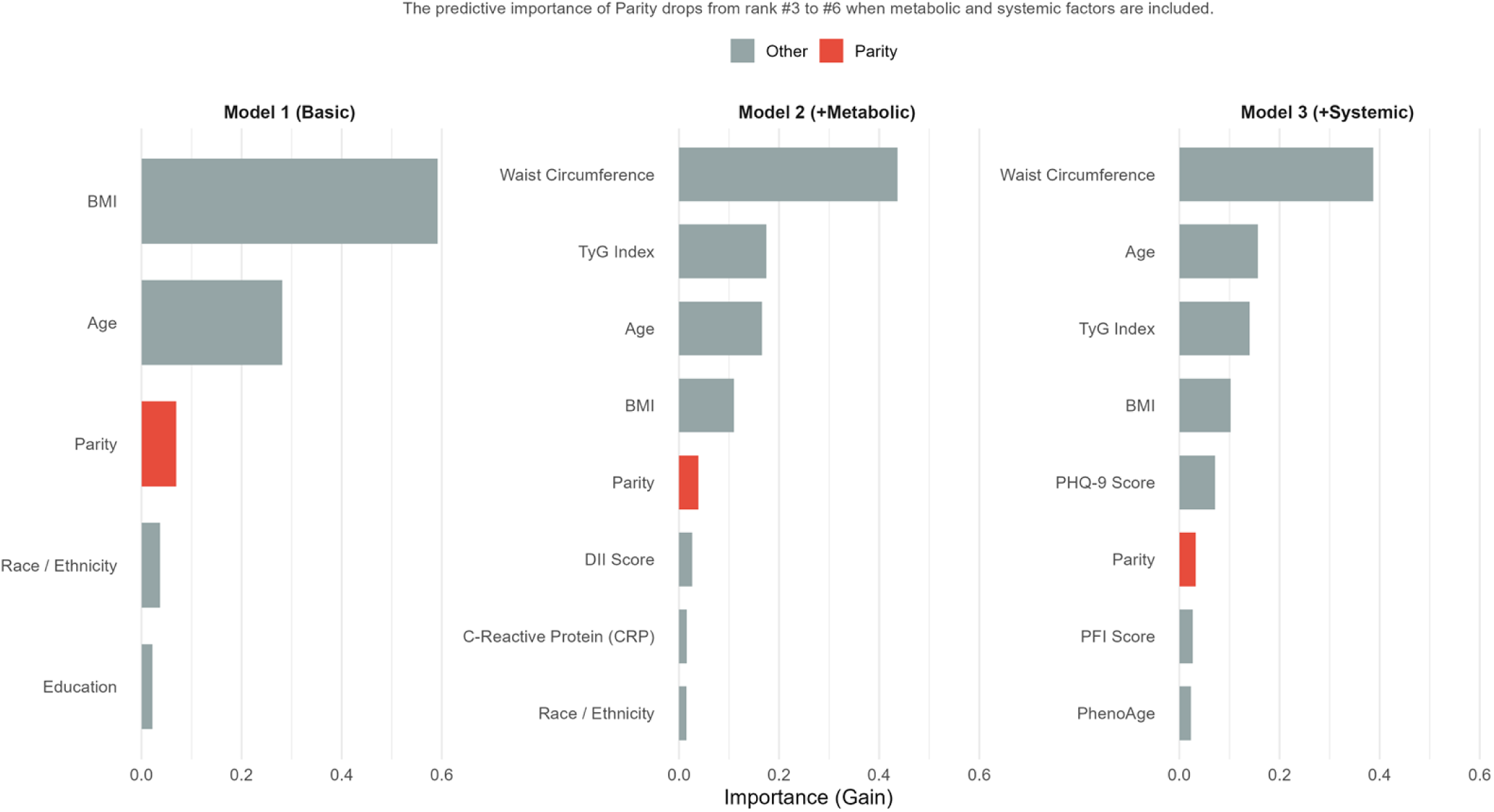
Feature importance shifts across nested models.

## 4. Discussion Principal Findings

Utilizing a nationally representative sample and an unsupervised machine learning approach, this study is the first to reveal that female pelvic floor dysfunction (PFD) is not a single clinical entity but can be deconstructed into at least two clinical subtypes with distinct biological profiles. This finding challenges the conventional understanding of PFD and provides a new theoretical framework for precision medicine in this field. Our primary discovery is the identification of a large “Metabolic-Inflammatory” PFD subtype, driven by central obesity and systemic metabolic dysregulation, which coexists alongside the classic subtype associated with age and parity.

## The Metabolic-Inflammatory Subtype: PFD as a Local Manifestation of Systemic Disease

The core finding of this study is the identification of Phenotype 1, the “Metabolic-Inflammatory” subtype. Patients in this group present with a severe metabolic syndrome profile, including exceptionally high BMI and waist circumference, a diabetes prevalence exceeding 20%, and a hypertension prevalence of 50%. Feature importance analysis further confirmed that waist circumference is the single most decisive factor in differentiating the subtypes, with a contribution far surpassing all other variables. This strongly suggests that for these women, PFD may not be an isolated structural issue of the pelvic floor but should rather be viewed as a local manifestation of a systemic metabolic disease. This perspective aligns with the emerging concepts of “meta-inflammation” and “inflammaging,” which posit that obesity-driven, chronic low-grade inflammation systemically damages vasculature, nerves, and connective tissue. The pelvic floor, as a region subject to significant mechanical stress and structural complexity, may be a particularly vulnerable target in this systemic pathological process (8,10,23–25).

Our validation analysis in the periodontitis sub-cohort provides strong external evidence for this hypothesis. We found that the prevalence of periodontitis was significantly higher in the PFD-positive group. Given that periodontitis is a well-established source and marker of systemic chronic inflammation, this association suggests that the pathophysiology of PFD may share common inflammatory pathways originating from the oral cavity or elsewhere. Conversely, we did not find a significant association between PFD and grip strength in our analysis of that sub-cohort, implying that the systemic factors driving PFD may be more closely aligned with the “metabolic-inflammatory” pathway than with a “sarcopenia/frailty” pathway.

## The Metabolically-Healthy Subtype: A Reaffirmation of Traditional Etiology

In stark contrast to Phenotype 1, Phenotype 2—the “Metabolically-Healthy” subtype—exhibited a metabolic profile nearly indistinguishable from that of the healthy controls. These women, however, were older and had a similarly high proportion of vaginal deliveries. The existence of this subtype clearly reaffirms that age-related tissue degeneration and delivery-associated mechanical injury represent an independent and robust traditional pathway to PFD. The value of our study lies in its ability to objectively distinguish this “classic” pathway from the newly identified “metabolic” pathway through a data-driven approach, demonstrating that there is more than one road to PFD.

## A New Perspective on the Role of Parity: The Mediation Hypothesis

Childbirth is widely recognized as one of the most significant initiating factors for PFD (26–28).However, our sensitivity analysis revealed a more complex picture. While parity ranked as the third most important predictor in a basic clinical model, its relative importance diminished significantly when metabolic and systemic health indicators were included. This finding suggests that the impact of parity on PFD may be largely mediated through its long-term effects on a woman’s metabolic health and systemic inflammatory state, rather than solely through direct mechanical injury from delivery. In other words, childbirth may act as a “trigger” that initiates a trajectory of long-term weight gain and metabolic dysregulation in susceptible women, with the latter ultimately becoming the core driver of PFD progression. This “mediation hypothesis” offers a new perspective for clinical practice: attention should be paid not only to parity itself but also to the long-term, holistic metabolic health management of postpartum women.

## Clinical Implications and Future Directions

The findings of this study have profound implications for the clinical practice and future research of PFD. First, they call for a shift from a “one-size-fits-all” treatment model to a “subtype-specific” precision medicine strategy. For patients with the “Metabolic-Inflammatory” subtype, the focus of treatment may need to extend beyond pelvic floor physical therapy or surgery to include aggressive weight management, improvement of insulin resistance, and control of systemic inflammation. Second, our predictive model demonstrates that these subtypes can be distinguished with high accuracy (AUC > 0.83) using only a few common clinical metrics (e.g., waist circumference, age, TyG index), laying the groundwork for the development of simple, practical clinical screening tools.

Future research should aim to validate the long-term evolution of these two subtypes and their differential responses to treatment in longitudinal cohorts. A critical unanswered question is whether the PFD symptom profiles (e.g., the prevalence and severity of SUI, UUI, and POP) differ between the subtypes, which will be key to refining subtype-specific treatment strategies. Furthermore, identifying specific biomarkers (such as particular inflammatory cytokines or metabolites) associated with each subtype will be the ultimate goal for achieving higher-precision diagnostics and developing targeted therapies.

## Strengths and Limitations

The primary strengths of this study are its foundation in the large, nationally representative NHANES database and its pioneering use of unsupervised machine learning to objectively deconstruct PFD heterogeneity. Our study design was rigorous, incorporating model validation, feature interpretation, and multiple sensitivity and validation analyses. However, several limitations must be acknowledged. First, the cross-sectional design of NHANES precludes the inference of causality. Second, some data relied on self-reporting, which may be subject to recall bias. Third, key objective validation metrics (e.g., periodontitis and grip strength) were available only in smaller sub-samples, limiting the generalizability of those specific conclusions. Finally, our analysis was based on a U.S. population, and its applicability to other ethnic and geographic groups requires external validation.

## 5. Conclusion

In conclusion, this study, through a data-driven approach, successfully deconstructs female pelvic floor dysfunction into two primary clinical subtypes: a “Metabolic-Inflammatory” type, primarily driven by central obesity and metabolic dysregulation, and a “Classic” type that follows the traditional pathway of aging and parturition-related injury. This finding reshapes our understanding of PFD pathophysiology and opens new avenues for developing personalized prevention and treatment strategies, marking a significant step toward an era of precision medicine for PFD.

## Data Availability

The data described in the manuscript were obtained from the National Health and Nutrition Examination Survey (NHANES), a publically available database. The data, code book, and analytic code are freely and publicly available without restriction at the Centers for Disease Control and Prevention website (https://wwwn.cdc.gov/nchs/nhanes/default.aspx).

